# Assessing the Performance of Machine Learning Models to Predict Neonatal Mortality Risk in Brazil, 2000-2016

**DOI:** 10.1101/2020.05.22.20109165

**Authors:** Luciana Correia Alves, Carlos Eduardo Beluzo, Natália Martins Arruda, Rodrigo Campos Bresan, Tiago Carvalho

## Abstract

Neonatal mortality figures are an important health’s problem, as the first month of life is the most vulnerable time for survival. Factors associated with neonatal mortality are complexly and influenced by the maternal and newborn biological characteristics, social conditions and the care provided by the health services. The aim of this study was investigated the association between features related and neonatal mortality risk in Brazil. Data came from two surveys: The Mortality Information System and Information System on Live Births. The final sample was composed of 302,943 children between 2006 and 2016. We highlight the proposition of a new approach based on machine learning to address the problem of neonatal mortality death risk classification. The results using three different machine learning classifiers points toward expressiveness of features, being newborn weight, Apgar at the first and fifth minute, congenital malformations, gestational weeks and number of prenatal appointments the six more expressive.

## Introduction

The decline in mortality is one of the greatest achievements of civilization. Unlike in developed countries, mortality declines in developing countries occurred at different time and rhythm (Palloni and Pinto Aguirre, 2011). As well as the case in Latin American countries, in Brazil, there was a significant improvement in life years after the 1930s. The importation of medical technologies associated with public health measures, such as immunizations, allowed for faster advances and in a short period.

In Brazil, mortality started declining, mostly at young ages, around 1940. Infant mortality decreased from 135 to 20 per thousand live births between 1950 and 2010. Between 1991 and 2010, the infant mortality rate dropped to 16.2 deaths per 1,000 live births and life expectancy at birth increased from about 50 to about 73 years over the same period (IBGE, 2010). The largest contribution to gains in life expectancy was due to falling infant mortality (Vasconcelos and Gomes, 2012). The changes in fertility have been even more remarkable, and with more dramatic implications. The average Brazilian woman had more than six children in the early 1960s and currently has less than two. Over time these changes in mortality and fertility alter the population age structure.

In 2014, about 7 million children below five years of age still died around the world. Of which, 3.4 million deaths occurred in sub-Saharan Africa, 2.3 million in South Asia, and fewer than 100,000 in the developed world. Globally, this death toll represents more than one child death every 5s. The vast majority of these children have died from diseases preventable or treatable with simple and low-cost medical techniques. Despite this depressing toll, tremendous progress has been achieved since the 1950s.

Worldwide, the proportion of children dying below five years of age declined from about 200 per 1,000 births around 1950 to 120 per 1,000 in 1980–85, and further to about 55 per 1,000 in 2005–10. Compared to the conditions of the 1950s, improvements in health care and sanitation have resulted in the survival of nearly 20 million children who would have died every year (Barbieri, 2015). In this transition process of mortality in Brazil, we can highlight the reduction in infant mortality for infectious and parasitic diseases, which are mainly risk factors associated with better life and sanitary conditions, hygiene, nutrition, access and care health.

Infant Mortality Rate (IMR) and Neonatal Mortality Rate (NMR) are an important measure of health in a population as a crude indicator of the poverty and socioeconomic level as availability and quality health services and medical technology in a specific region. A decrease in NMR and IMR results in the improvement of infant mortality and survival, which can positively influence the national public state of health (Chung et al., 2011). The neonatal mortality accounts to approximately 60% of the infant mortality in developing countries (Singha et al., 2016). This dimension of infant mortality is important because, for the World Health Organization (WHO, 2018) and United Nations Children’s Fund (UNICEF, 2015), the first month of life is the period which the child is more vulnerable.

Infant Mortality is a worldwide concern in public health as defined by the United Nation (UN) as the global development goals when setting as target the reduction of the infant mortality until 2015. Brazil achieved this Millennium Development Goal, but national rates do not reveal the persistent inequalities remaining between geographic regions and population groups. Regions and populations with lower incomes are at greater risk of infant death. In addition to the disparities arising from socioeconomic and geographic factors, infants in the first week of life (early neonatal death) did not reduce satisfactorily and now represent the greatest challenge to the advancement of addressing infant mortality in the country (Ministry of Health, 2015).

The problem of infant mortality in Brazil has become relevant, since the available data and their respective analyzes point out to the persistence of disparities between regions, states and populations with different socioeconomic characteristics, despite the constant tendency of general decline (Ministry of Health, 2015).

Developed countries have on average four neonatal deaths per 1,000 live births and, in Brazil, we had a neonatal mortality rate in 2017 of approximately nine deaths per 1,000 live births. Since the enactment of the Federal Constitution of 1988, a large part of the burden of coping with neonatal mortality has been imposed on municipalities, which have taken on a prominent position in the implementation of public health policies (Machado et al., 2015).

In 2003, Mosley and Chen proposed a hierarchical model based on the hypothesis that socioeconomic factors determine behaviors, which, in turn, have an impact on a set of biological factors. According to their model, biological factors are those directly responsible for the death. The hierarchical model brings a great advance to the development of public policies, since information coming from studies that are limited to only a group of risk factors result in inadequate recommendations to assess the deaths among children, as they present a limited vision of the phenomenon. Therefore, factors associated with neonatal mortality are complexly articulated and influenced by the maternal and newborn biological characteristics, social conditions and the care provided by the health services (Nascimento et al., 2012; França and Lansky, 2016). We believe that different characteristics of the mother and the newborn as maternal obstetrics, related to the newborn and related to care assistance on prenatal and delivery can predict neonatal mortality more than socioeconomic characteristics of the mother.

There has been growth in the volume of demographic and epidemiology studies in Brazil that explore the connection between specific factors related to infant and neonatal mortality, but that use traditional regression models (Duarte and Mendonça, 2005; Nascimento et al., 2012; Lima et al., 2012; Migoto et al., 2018; Garcia et al., 2019). It is important to recognize, therefore, the need for the use of specialized tools to increase the power of the studies with the aim of to allow the visualization and manipulation of data from large population segments, thus enabling the formulation of follow-up indicators. Neonatal mortality is a complex phenomenon, involving interactions of several characteristics and requiring a large volume of data for its full understanding. Machine learning methods have great potential for a better understanding of the interactions between different factors but are rarely used in neonatal mortality studies in Brazil. Thus, the application of machine learning in this context is innovative to the Brazilian reality. With models that make such analyses more efficient and effective, in addition to preventing neonatal deaths, it is also intended to improve the care provided to women, as there is frailty in the integration between prenatal care and delivery care.

The aim of this study was to predict risk of neonatal death and to assess the feature importance based on machine learning approaches between 2006 and 2016 in Brazil. The hypothesis of the present research is that neonatal mortality is a complex phenomenon, involving interactions of several characteristics and requiring a large volume of data for its full understanding. In this sense, we believe that traditional regression models may not be enough to understand this problem, since the assumptions of parametric modelling are unrealistic for investigations of this nature.

## Material and Methods

The present study is an observational, retrospective cohort study based on secondary data of births and deaths of infants related with this cohort in Brazil between 2006 and 2016. Data came from by *Sistema de Informação sobre Mortalidade* (SIM — Mortality Information System) and *Sistema de Informação sobre Nascidos Vivos* (SINASC-Live Birth Information System) from DATASUS (Health Informatics Department of the Brazilian Ministry of Health). To identify the deaths related with the cohort, the Federal Infant and Fetal Death Investigation Module was used, which automatically pairs the infant death declarations (DD) and their respective birth declarations (BD), based on the BD number.

Figure 1 illustrates an overview of the process to linkage the data from SIM with data from SINASC and data cleaning aiming at having characteristics of the newborn, demographic and socioeconomic characteristics of the mother and whether the newborn died before the 28th day of life or not. The linkage technique was used to relate the datasets through the application of the deterministic method, that is, we use the common variable for both systems, Number of Live Birth Statement (NUMERODN).

**Figure 1:**
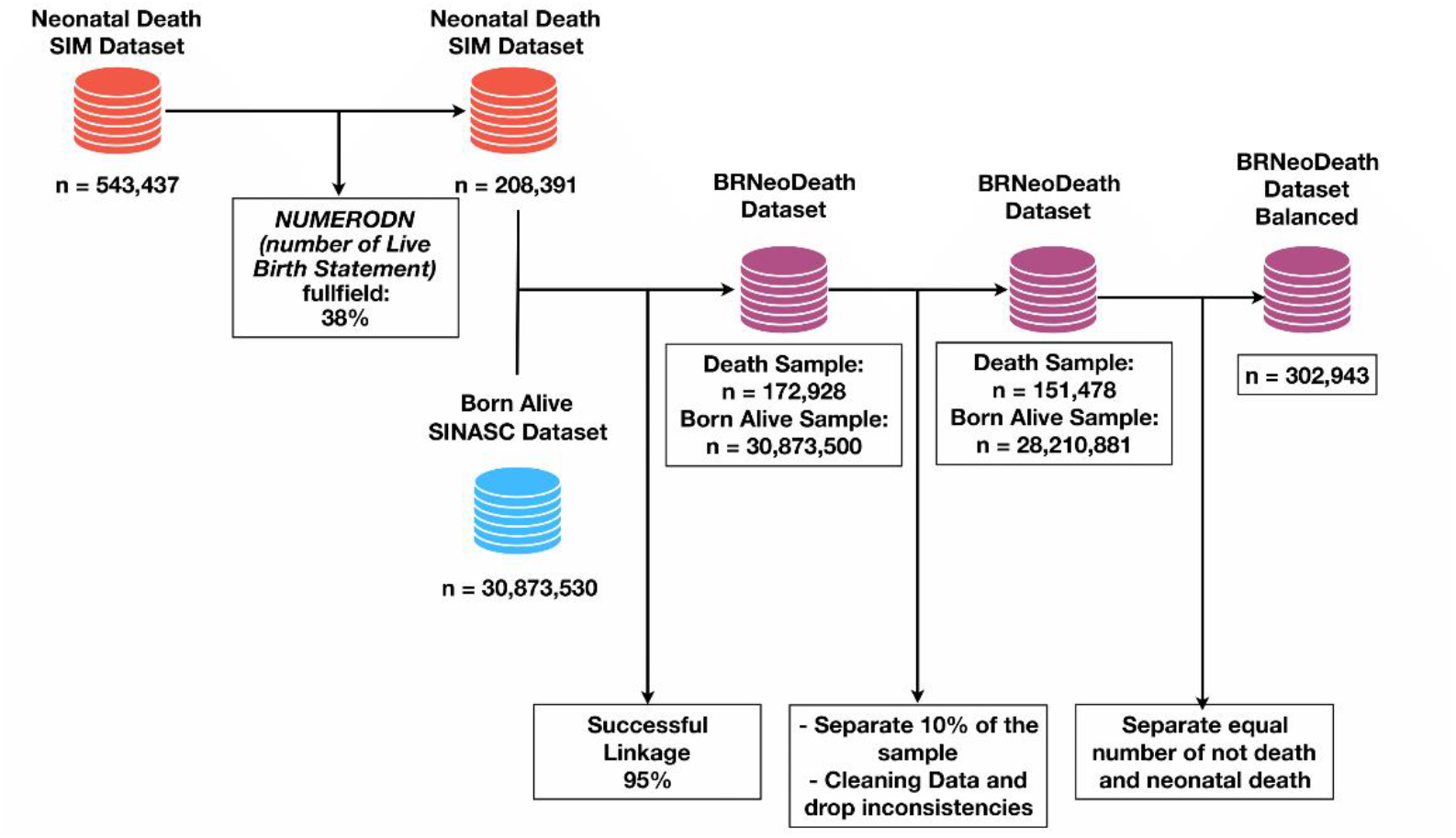
Flowchart of the process to linkage the data from SIM and SINASC for balanced dataset

In order successfully link these datasets it was necessary that NUMERODN field be completed in the death certificate and, despite the fact that it is mandatory to fill this field in the deaths up to one year of age, only 38% were fulfilled (n=208,391). From this percentage, it was possible to join the two datasets in 95% of the cases resulting in a large dataset that we call BRNeoDeath with initially 30,873,500 observations.

After the process of linkage, from our entire dataset, we keep a sub-sample of 10% randomly selected data (respecting classes distribution) without doing any preprocessor. This kind of procedure is strongly applied in challenges involving machine learning and data driven approaches and it is performed to allow a fair comparison between different methods proposed to solve the same problem. With the other 90%, we applied a data cleaning to remove inconsistencies such as duplicate observations and categories not in the data dictionary. It was chosen for these cases to exclude the observation. Besides that, to take care of the missing data we perform two different approaches. For variables initially continuous, such as weight and mother age, we use the mean of that given column while. For variables categorical, we used the most frequent value of that given category resulting in a dataset with a sample of neonatal death of 151,473 observations and 28,210,886 of born alive.

As we can see, the BRNeoDeath have unbalanced class distribution, where the percentage of death class samples are outnumbered by the percentage of living class samples, being 99.4% of the living class, and just 0.6% of the dead class. This problem is often referred to in the literature as the ‘class imbalance’ and can generate low performance classifiers, especially when predicting low (minority) represented classes (Prati et al., 2009). Given the nature of unbalanced datasets, we use a sub-sample from BRNeoDeath. That sub-sampled dataset consists of all the positive samples (death class) and the same amount of negative samples (alive class) randomly selected across the entire dataset resulting in a sub-sample of 302,943 observations, representing the final sample.

For the present investigation, the variables were divided by: (1) demographic and socioeconomic maternal conditions: maternal age (8-14, 15-19, 20-24, 25-29, 30-34, 35-39, 40-44, 45-49, 50 or more years), maternal education (0, 1-3, 4-7, 8-11, 12 or more), marital status (single, married/stable relationship, widowed, separated/divorced) and race/skin color (white, black/brown, yellow, indigenous); (2) maternal obstetrics: number of live births (0-3, 4-7, 8-10, 11 or more live births), fetal losses (0 - 3, 4 - 7, 8 - 10, 11 or more), number of previous gestation (0-3, 4-7, 8-10, 11-14, 15 or more), number of normal labor (0-3, 4-7, 8-10, 11-14, 15 or more) and number of cesarean labor (0-3, 4-7, 8-10, 11-14, 15 or more), type of pregnancy (single, double, triple or more); (3) related to the newborn: newborn weight (< 2,500; 2,500-2,999; 3,000-3,999; 4,000 or more grams-g), gestational weeks (< 22, 22-27, 28-31, 32-36, 37-41, 42 or more weeks), Apgar at the first minute (0 - 3: severe, 4 - 6: moderate, 7: light, 8-10: optimum), Apgar at the fifth minute (0 - 3: severe, 4 - 6: moderate, 7: light, 8-10: optimum), and congenital malformations (yes, no) and type of presentation of the newborn (cephalic, podalic or breech, shoulder); (4) related to previous care: number of prenatal appointments (0, 1-3, 4-6, 7 or more), labor type (vaginal, cesarean section), cesarean section occurred before labor (yes, no, not apply), labor induced (yes, no), childbirth care (doctor, nurse, midwife, other) and Robson 10-groups classification. Robson 10-groups is a measure of cesarean rate assessment and monitoring. This classification distributes women into 10 groups bases on five characteristics: early labor (spontaneous, induced or cesarean), gestational age, fetal presentation, number of fetuses and parity (nulliparous, multiparous with and without previous cesarean section). The Robson 10-groups was divided: Group 1: Nulliparous with single cephalic pregnancy, greater than 37 weeks of gestation in spontaneous labour; Group 2: Nulliparous with single cephalic pregnancy, greater than 37 weeks gestation who either had labour induced or were delivered by caesarean section before labour; Group 3: Multiparous without a previous uterine scar with single cephalic pregnancy, greater than 37 weeks gestation in spontaneous labour; Group 4: Multiparous without a previous uterine scar, with single cephalic pregnancy, greater than 37 weeks gestation who either had labour induced or were delivered by caesarean section before labour; Group 5: All multiparous with at least one previous uterine scar, with single cephalic pregnancy, greater 37 weeks gestation; Group 6: All nulliparous women with a single breech pregnancy; Group 7: All multiparous women with a single breech pregnancy, including women with previous uterine scars; Group 8: All women with multiple pregnancies, including women with previous uterine scars; Group 9: All women with a single pregnancy with a transverse or oblique, including women with previous uterine scars and Group 10: All women with a single cephalic pregnancy less than 37 weeks gestation, including women with previous scars.

Descriptive statistics were calculated for all variables. In order to analyze the main determinants of the neonatal death risk in Brazil, the present study used the Machine Learning models. Three algorithms were tested and analyzed: Random Forest, Extreme Gradient Boosted Trees (XGBoost) and Support Vector Machine. The algorithms used were able to classify the determinants, highlighting the most powerful determinants.

Machine learning models were used across one round of experiments using different metrics to measure to identify its effectiveness in neonatal death risk classification task.

### Machine Learning Methods

The Machine Learning method is useful as a replacement or complement to parametric regression. Unlike traditional regression-based approaches, machine learning does not impose a parametric model linking a dependent variable with independent variables. The key idea is to let the algorithm find the path to the result and links between the independent variables. This way it is possible to automatically look for relationships and interactions between the independent variables. Moreover, collinearity and assumption violations are not important concerns depending on the chosen algorithm (De Rose and Pallara, 1997; Billari et al., 2006).

There are two broad categories of Machine Learning techniques, ‘supervised’ learning and ‘unsupervised’ learning. ‘Unsupervised learning’ focuses on methods for finding patterns in data and for data reduction (Kuhn and Johnson, 2013). Unsupervised learning is widely used to group unlabeled data based on the similarity of characteristics, while supervised learning is appropriate for predictive modelling by constructing some relationships between socioeconomic characteristics, childbirth (as inputs) and the outcome of interest (as a result of this study, neonatal mortality). The present analysis used supervised learning in a balanced dataset.

Although the primary purpose of these techniques is to build a predictive model, these methods can usefully be used to examine how a (potentially large) set of independent variables is linked to an outcome. Therefore, these techniques can be used as a nonparametric alternative to regression-type approaches by studying the relationships between a set of independent variables and a dependent variable (Kuhn and Johnson, 2013).

An individual feature of supervised algorithms is that the building model is data-driven so that they can automatically adjust to complex relationships, overcoming mainly variable selection and model building efforts. More specifically, Machine Learning algorithms can automatically detect nonlinearities and no additives. These algorithms can be useful for improving data analysis because of their flexibility, particularly when dealing with large data sets (in terms of sample size and the number of covariates).

### Data preprocessing

Transformations of predictor variables may be required. Some modelling techniques may have strict requirements, such as predictors having a common scale. Most data sets require some degree of preprocessing to expand the universe of possible predictive models and optimize the predictive performance of each model (Kuhn and Johnson, 2013). In the present study, all variables that were continuous were transformed into categorical (maternal age, Apgar at the first minute, Apgar at the fifth minute, number of live births, fetal losses, number of the previous gestation, number of normal labor and number of cesarean labor). So it was possible to transform all the variables so that the machine learning models could interpret the categories of each variable ordinally presented, the hierarchical relationship between categories, that means that the model could understand that the higher the category value is, more effective it is.

The transformation adopted was that of one hot encoding, that is the process by which all categorical variables were transformed into dummy variables. This process generates a vector of N positions for each existing feature, where N is given by the number of unique values (different categories in that feature). Position matching feature category is filled with 1 while rest of positions is filled with 0. For example, the marital status variable has four categories, each of these categories turned into a column where each position was filled with zero and one depending on the occurrence of that category. Data were separated by 80% for training and 20% for testing.

### Metrics

From the confusion matrix illustrated below (Figure 2), it was possible to calculate the main measures that were used for the comparison of the algorithms. Each algorithm generated the confusion matrix values. Rows are the actual classes and columns are the predicted classes. The diagonal values will be the values that the model generated as correct predictions.

**Figure 2:**
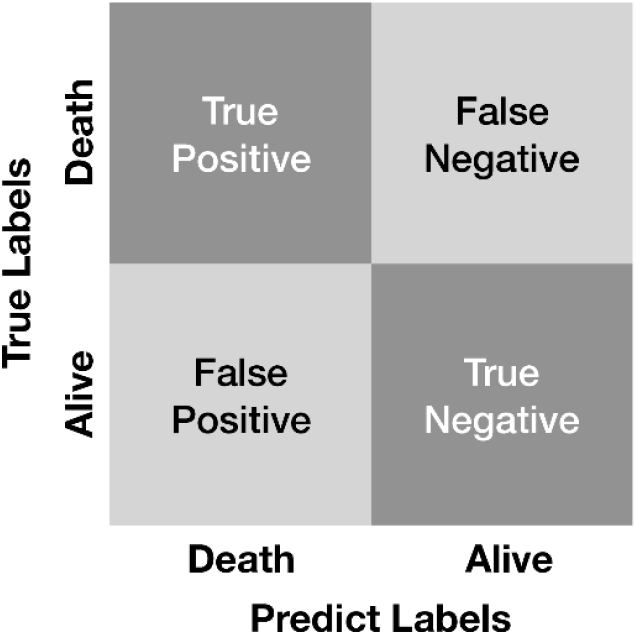
Confusion matrix for estimating metrics

The following metrics were calculated: (1) accuracy: determines the number of predictions made correctly by the model overall predictions made; (2) sensitivity: ratio of true positives to true positives plus false negatives; and (3) specificity: ratio of true negatives to true negatives plus false positives.

Besides that, we use as metric the Receiver Operating Characteristic Curve (ROC Curve) that is a graphical representation that allows to measure behavior of a binary classifier system as its discrimination threshold is varied. Y-axis report True Positive Rate - TPR (also named Sensitivity) while X-axis report True Negative Rate - TNR (also named Specificity). Area below the ROC curve (graph with sensitivity on one axis and specificity on the other) indicates the choice of the cutoff point for the best combination of sensitivity and specificity measurement. Area Under a Curve (AUC) represents the integral under ROC Curve values.

### Models Construction

The last step of the proposed method consists in construct and evaluation the performance of three machine learning algorithms (support vector machine, random forests, XGBoost) to classify samples according to its death risk. Chosen given their good results achieved on health problems.

Support Vector Machines (SVM) (Cortes, 1995), is one of the most common methods applied on supervised classification problems mainly because it’s excellent accuracy and generalization properties (Podda et al., 2018; Hsieh et al., 2018). The basic concept behind SVM consists in finding a hyper-plane that can separate data according to their classes. To accomplish this task the method projects features into an M dimensional space using kernel application.

Tree-based methods provide easy comprehension of their outcomes, as well as the interaction between features used for classification. Random Forests are a specific kind of tree-based method that generates multiple trees with a random subset of features for training and testing, leading to higher diversity and more robust predictions (Breiman, 2001). This kind of approach has a good performance in different literature problems, including prediction in different contexts of child mortality using features related with child and mother (Nguyen, 2016; Pan, 2017; Podda et al., 2018).

XGBoost has been proposed to push the limits of processing power for boosted trees algorithms. These techniques have been refined to extract most of the system hardware in order to provide a high-quality model. The method explores a sparsity-aware algorithm for sparse data and weighted quantile sketch for approximate tree learning (Chen and Guestrin, 2016). A variant of this model has been used in Podda et al. (2018), the Gradient Boosting Machine (GBM), presenting good performance on predicting preterm infant survivor.

The methods were implemented using Python programming language (3.6), along with the Scikit-Learn (0.21.2), XGBoost (0.90), Pandas (0.24.2), MatplotLib (3.1) and Shap (0.29.3) libraries. All the experiments were performed using a machine with 40 CPU cores, 4 GPU TitanX 12 GB, 120 GB of RAM and 8 TB of storage, running Ubuntu 18.04 (64 bits).

## Results

In Brazil, as shown in Figure 3, the Infant Mortality Rate has decreased over the years. However, the Neonatal Mortality Rate has represented a higher proportion of Infant Mortality cases. Between 2006 and 2016, the infant mortality rate dropped to approximately 13 deaths per 1,000 live births. The neonatal mortality rate decreased from 11 deaths to about 9 per 1,000 live births in the same period.

**Figure 3.**
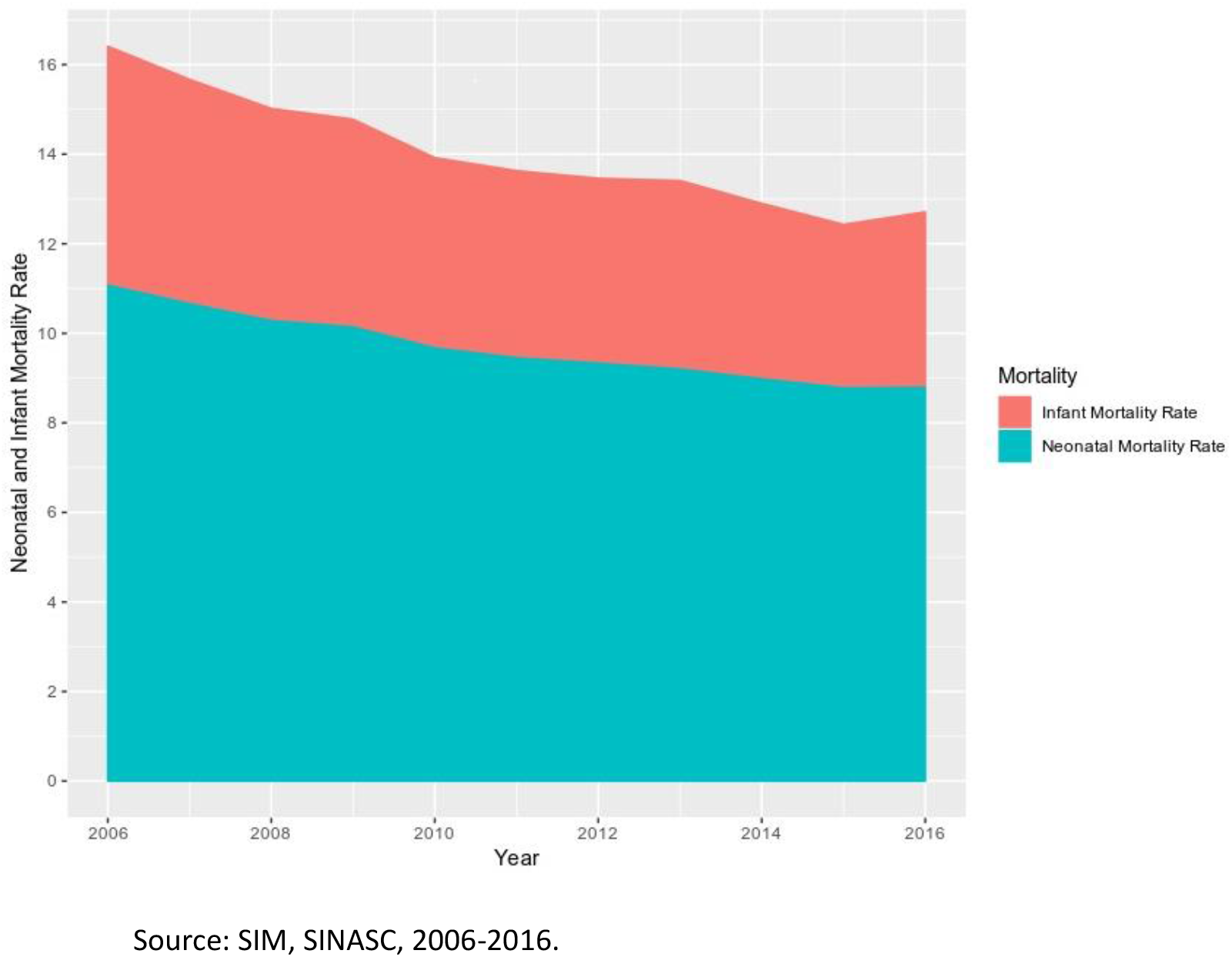
Participation of the Neonatal Mortality in the Infant Mortality, Brazil, 2006 – 2016

Table 1 displays the characteristics of the sample. Most mothers were between 15 and 29 years old, but mother’s age was predominantly 20 to 24 years old (25.95%); 50.88% of the mothers had 8 to 11 years of education; 54.80% were either married or in a stable relationship; 64.61% were black/brown. Most mothers had 0 to three children (96.73%). About 99.85%, 97.92%, 98.70%, and 99.52% had 0 to three fetal losses, previous gestation, normal labor and cesarean labor, respectively. Approximately 93.81% had a single pregnancy; 40.49% of the newborns were born weighing less than 2,500 grams and 38.80% had 3,000 to 3,999 grams; 60.99% had 37 to 41 gestational weeks. Most babies scored eight to 10 on Apgar in the first and fifth minutes (29.16 and 60.86, respectively).

**Table 1:**
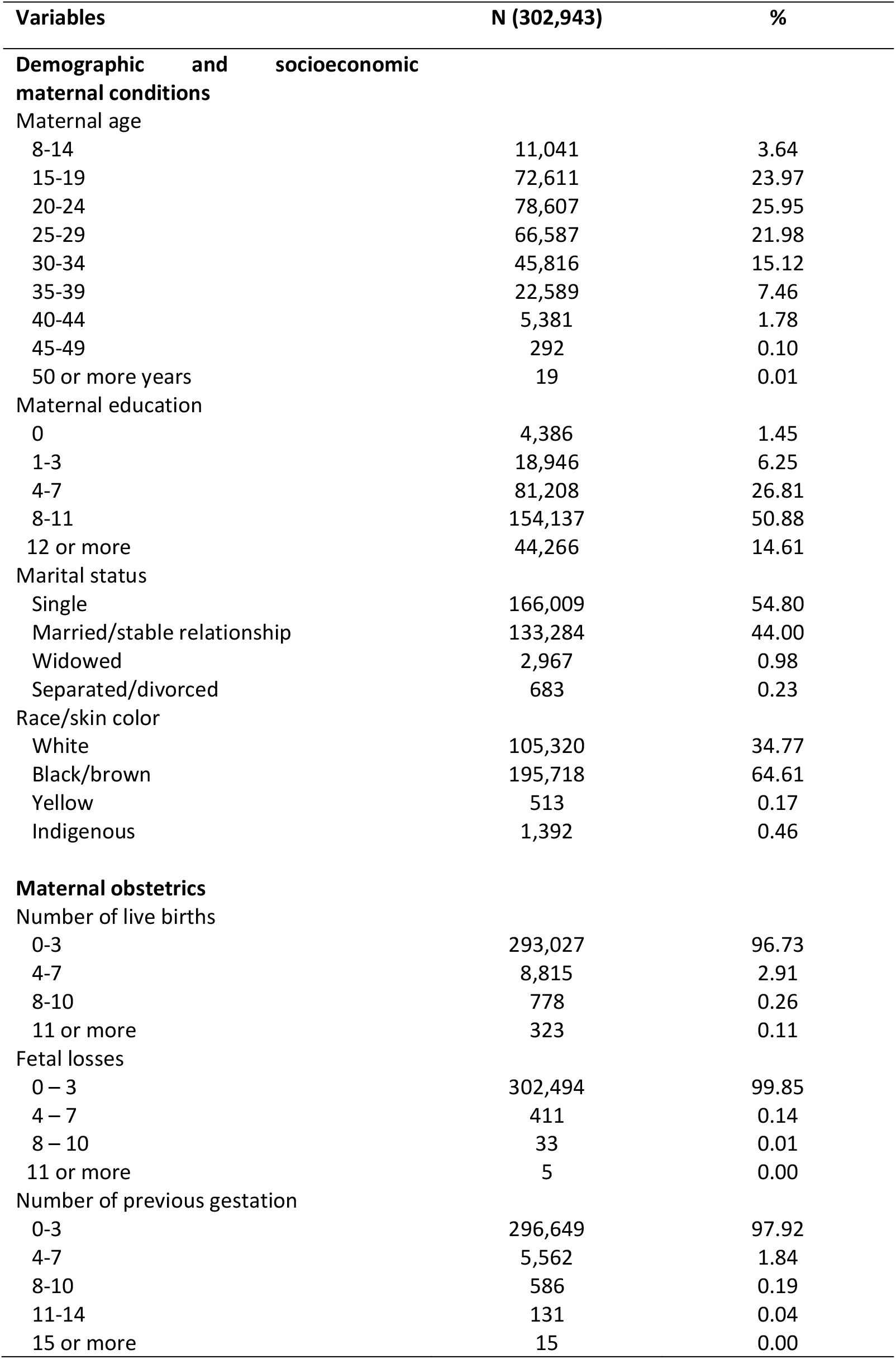
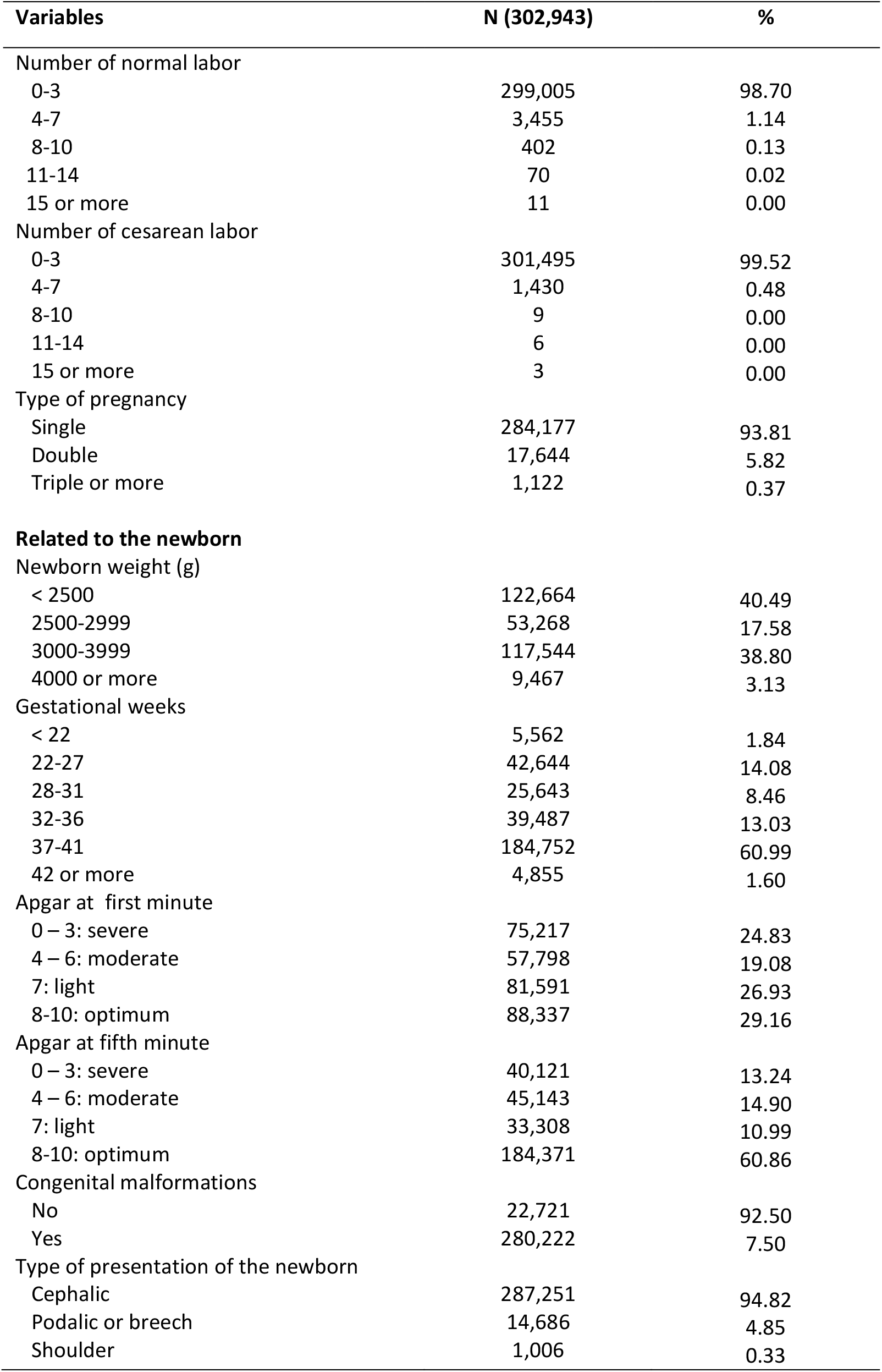
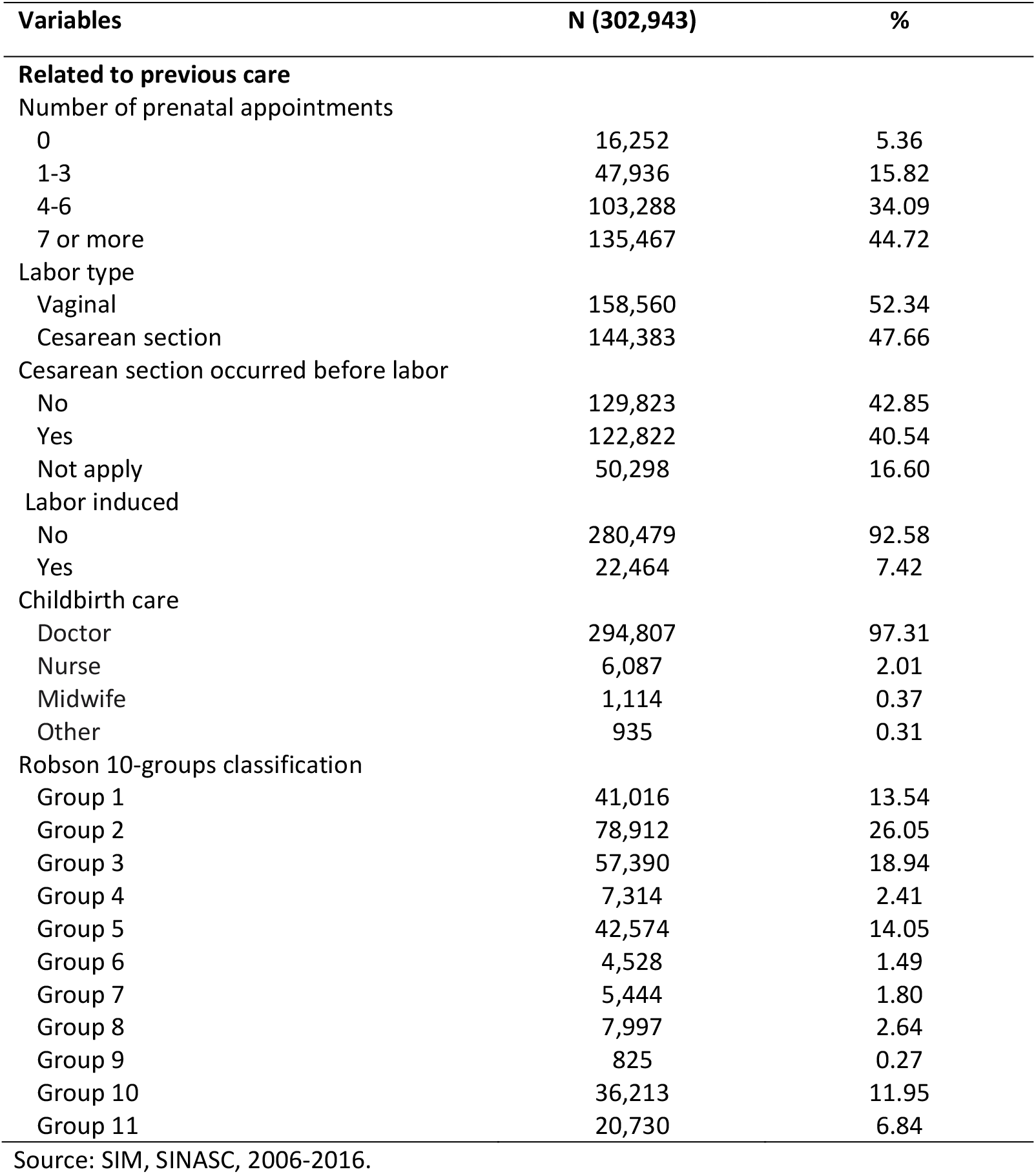
Relative Distribution (%) of demographic, socioeconomic, maternal obstetrics, related to newborn and previous care characteristics in Brazil, 2006-2016.

About 44.72% had seven or more prenatal appointments; 52.34% vaginal labor; 97.31% childbirth care for the doctor; and 26.05% were Robson Classification Group 2.

Approximately 53.8% of the men babies died in the period. Figure 4(a) and Figure 4(b) showed the number of days until death in neonatal death cases for the total and sex, respectively. Figure 4(a), point toward death happening until 6^th^ day after delivery in 75% of samples. The median was until 2 days after birth. For sex, this analysis showed toward death happening until 2 days for 50% and 6^th^ day after delivery in 75% of samples for both sexes.

**Figure 4(a).**
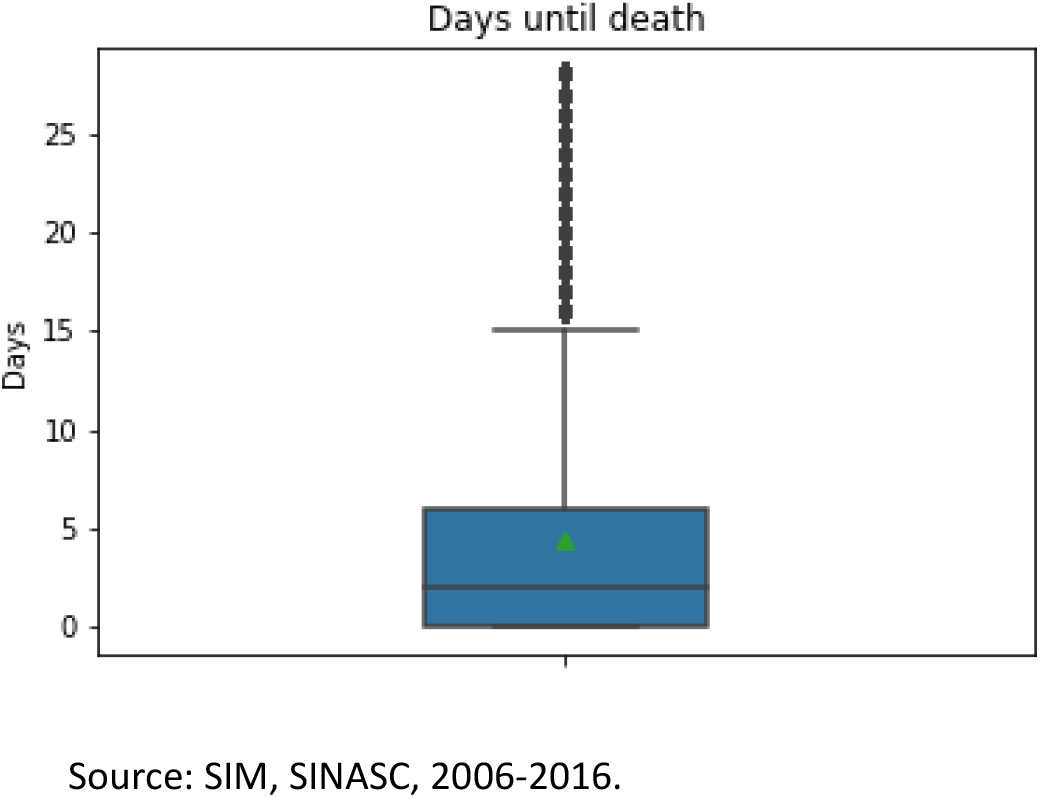
Days until Death of the Neonatal Mortality, Brazil, 2006 - 2016

**Figure 4(b).**
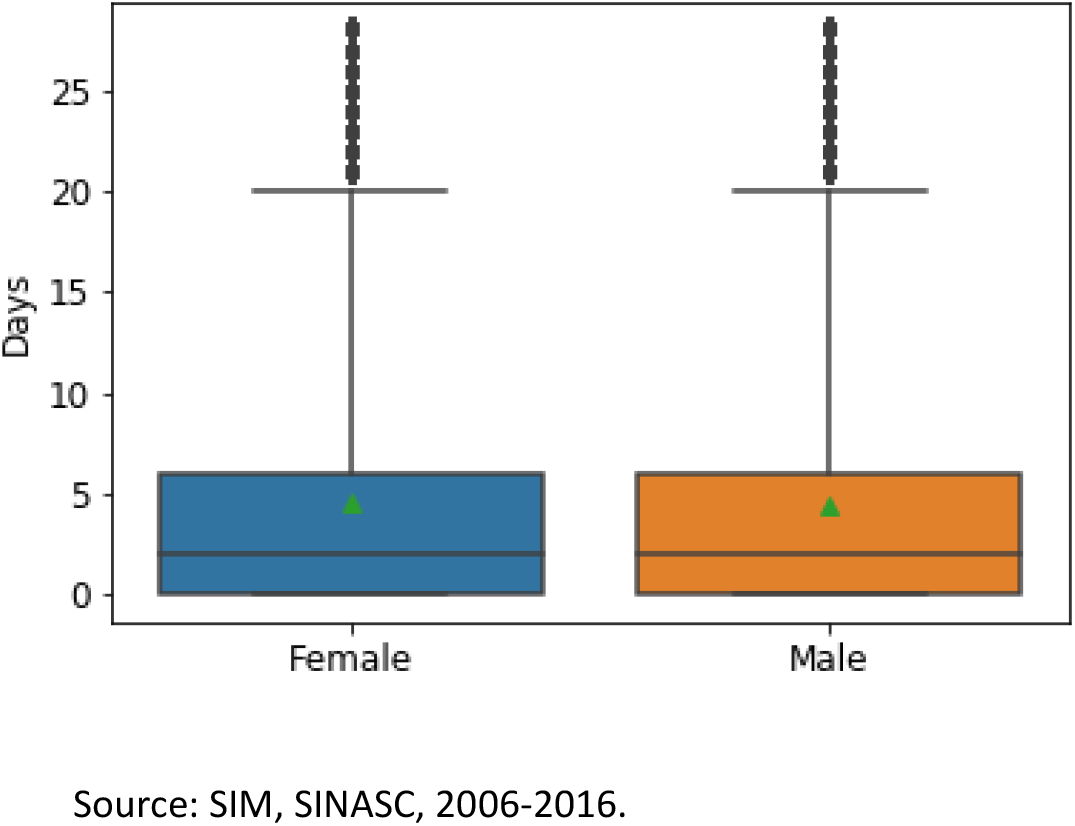
Days until Death of the Neonatal Mortality by sex, Brazil, 2006 - 2016

When analyzing feature newborn weight, 72.70% of newborns who died before the 28 days of life (death class) had insufficient weight - below 2,500 grams, whereas in the alive class only 8.30% were underweight. In relation to maternal age, in death class, that more than 35 years old showed higher neonatal mortality compared to younger mothers. Mothers with 12 or more years of education and with 7 or more prenatal appointments experienced lower neonatal mortality (13.1% and 29.4%, respectively).

Furthermore, results for gestational weeks showed that 47.7% of death class occurred below 31 weeks of gestation, had a severe or moderately severe evaluation, while 91.5% of the alive class had the mothers had more than 37 weeks of gestation (Table2).

**Table 2:** Relative Distribution (%) of balanced class distribution for death and alive according selected characteristics in Brazil, 2006-2016.

| Variables | Alive (%) | Death (%) |
| --- | --- | --- |
| Maternal age |  |  |
| 8-14 | 2.8 | 4.5 |
| 15-19 | 22.9 | 25.0 |
| 20-24 | 27.2 | 24.7 |
| 25-29 | 23.3 | 20.6 |
| 30-34 | 15.5 | 14.7 |
| 35-39 | 6.9 | 8.1 |
| 40-44 | 1.4 | 2.2 |
| 45-49 | 0.1 | 0.1 |
| 50 or more years | 0.0 | 0.0 |
| Maternal education |  |  |
| 0 | 1.3 | 1.6 |
| 1-3 | 6.3 | 6.3 |
| 4-7 | 26.3 | 27.3 |
| 8-11 | 50.1 | 51.7 |
| 12 or more | 16.1 | 13.1 |
| Number of prenatal appointments |  |  |
| 0 | 2.3 | 8.5 |
| 1-3 | 7.8 | 23.9 |
| 4-6 | 29.9 | 38.3 |
| 7 or more | 60.1 | 29.4 |
| Newborn weight (g) |  |  |
| < 2500 | 8.3 | 72.7 |
| 2500-2999 | 24.1 | 11.1 |
| 3000-3999 | 63.0 | 14.6 |
| 4000 or more | 4.6 | 1.6 |
| Gestacional weeks |  |  |
| < 22 | 0.0 | 3.6 |
| 22-27 | 0.3 | 27.9 |
| 28-31 | 0.7 | 16.2 |
| 32-36 | 7.5 | 18.6 |
| 37-41 | 89.4 | 32.6 |
| 42 or more | 2.1 | 1.2 |

Source: SIM, SINASC, 2006-2016.

We evaluate the proposed method on a dataset and an average ROC curve for each machine learning method. All evaluated methods present a similar performance on average, with ROC curves almost overlapped and an AUC of 93.93%, 92.56% and 92.43% for XGBoost, Random Forests (RF) and Support Vector Machine (SVM), respectively (Figure 5). In Figure 6, we depict the confusion matrix for all evaluated classifiers at the best threshold point of the ROC Curve. Accuracy reported on optimal ROC curves for Random Forest, Support Vector Machine and XGBoost are 87%, 88% and 89%, respectively.

**Figure 5.**
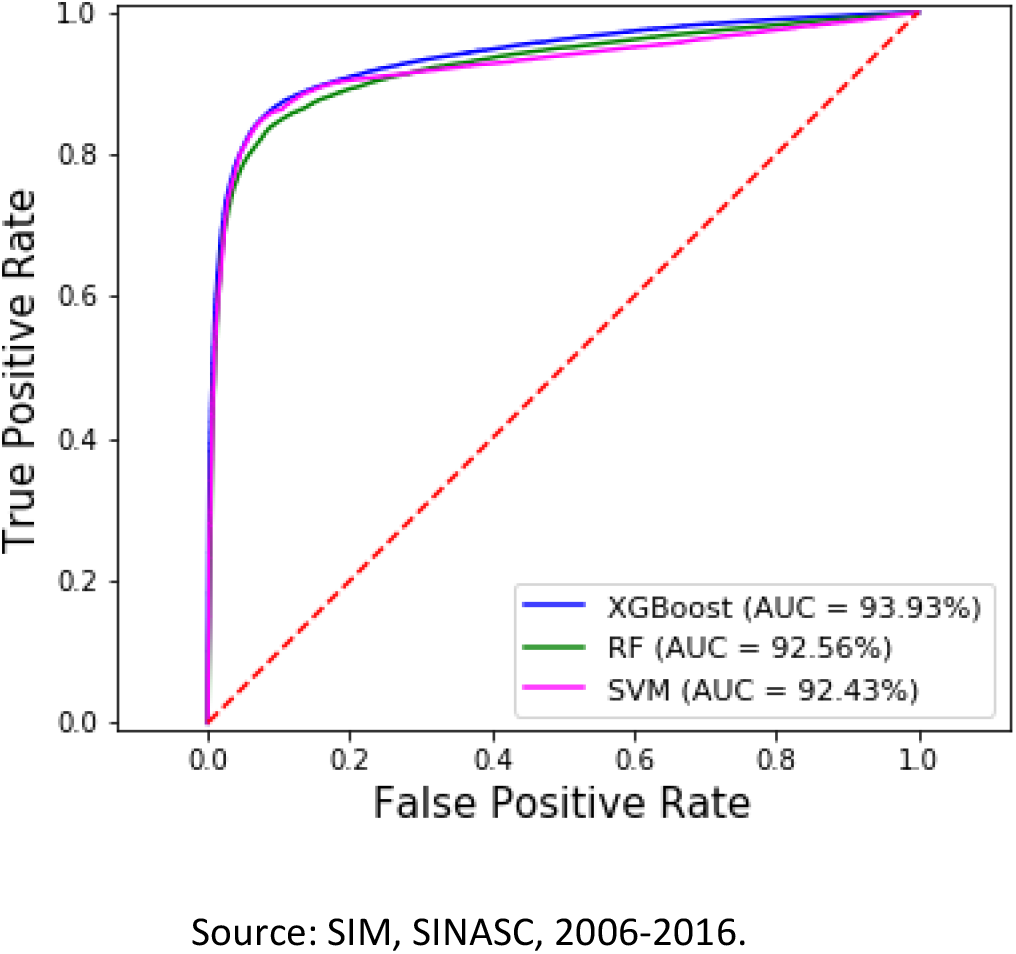
ROC Curve for all the evaluated models

**Figure 6.**
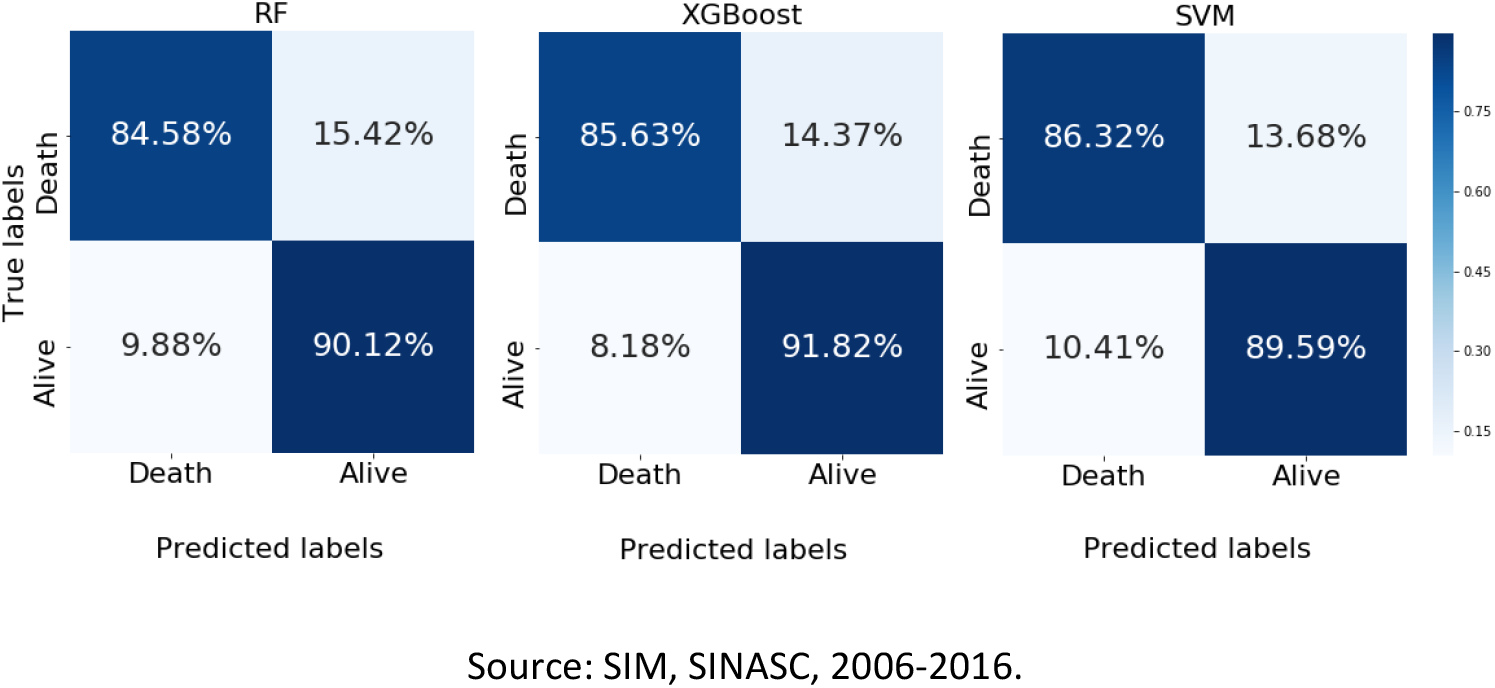
Confusion matrix at the optimal ROC Curve point for evaluated classifiers

The feature importance is a measurement that can point out the features with major relevance in our models. Using an XGBoost model, which was the model with the best performance, the results showed that the variables presenting a higher degree of importance (highly correlated with label class) were the newborn weight, Apgar at the fifth minute, congenital malformations, Apgar at first minute, gestational weeks and number of prenatal appointments. Maternal characteristics was not important (Figure 7).

**Figure 7.**
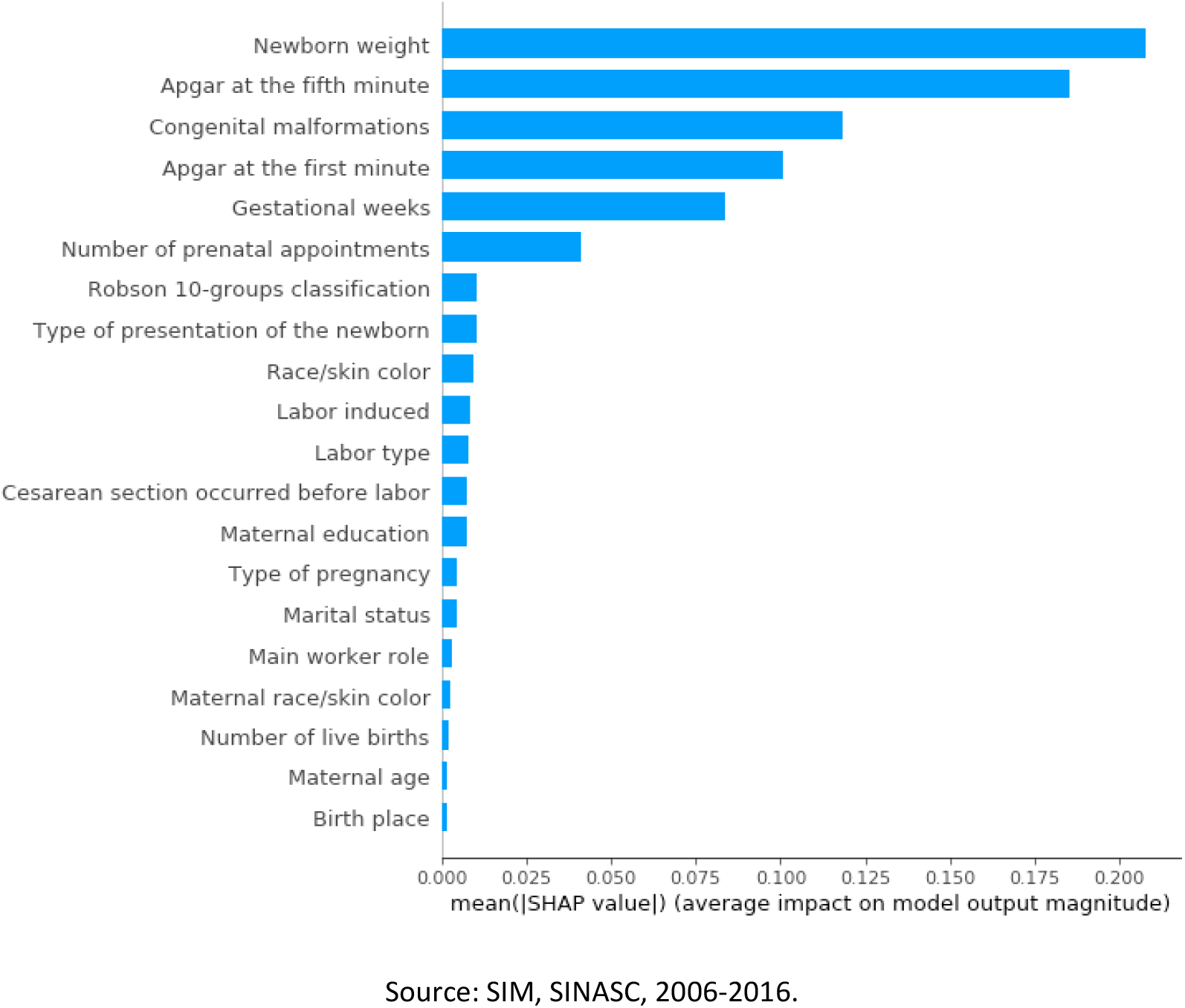
Feature Importance for XGBoost model

## Discussion

The present study assessed the risk of neonatal death and the feature importance based on machine learning approaches between 2006 and 2016 in Brazil.

Although the infant mortality rate has decreased over the years, the results revealed the neonatal mortality rate has represented a higher proportion in infant mortality cases in the period analyzed. Other studies have shown similar results. Gaíva et al. (2013) found an increase in neonatal deaths and the decrease in post-neonatal deaths in the city of Cuiaba in 2010. Most of the neonatal deaths worldwide occur 75% in the first week of life and more than half happens in the first 24 hours (WHO, 2018). In Brazil, demographic and epidemiological studies indicates this behavior is not different, being 70% on the first week and more than 50% in the first 24 hours (França and Lansky, 2009). In poor countries of Africa, neonatal deaths account for slightly more than 30% of infant mortality, in view of the unfavorable living and health conditions of the population that elevate post-neonatal mortality (Waldemar et al., 2010).

Mortality in the first days of life express the complex conjunction of biological, socioeconomic and care factors related to the protection of pregnant women and newborns (Duarte, 2005). According to UNICEF, in global scale, 2.5 million children died in the first month of life in 2017 alone – approximately 7,000 neonatal deaths every day – most of which occurred in the first week, with about 1 million dying on the first day and close to 1 million dying within the next six days.

An interesting relationship investigated in the present study was represented by sex of dead newborns and the number of survived days. United Nations (2011) point toward some biological advantage in women in relation to newborn men. Women have less vulnerability to perinatal conditions congenital anomalies and lower respiratory, intestinal infections, presenting higher survival rates in the first year of life. Despite women present a lower percentage of death in the neonatal mortality rate, we found that the number of survived days was similar for men and women.

In fact, neonatal mortality is still a problem for Brazil, but reducing infant mortality reflects an increase of public investments on public health that happened in the last years.

Some demographic, socioeconomic, maternal, infant and health care assistance-related aspects contribute to these deaths. Machine learning models revealed that newborn weight, Apgar at the fifth minute, congenital malformations, Apgar at the first minute, gestational weeks and number of prenatal appointments were the six most relevant features for neonatal deaths in Brazil.

Approximately one-quarter (28%) all children worldwide are born with low birth weight (Lai et al., 2017) and roughly 60 and 80% of all neonatal deaths are associated with this factor (Nascimento et al., 2012). Low birth weight infants are more vulnerable to pulmonary immaturity problems and metabolic disorders, which may cause or aggravate some events that affect them, increasing the risk for mortality. Thus, low weight and poor ratings in Apgar 1 and 5 minutes are warnings for possible future complications in the child, creating an alert on the risk of this newborn dying in the first days of life. Furthermore, the study of Nascimento et al. (2012) identified greater neonatal mortality among premature and low birth weight. Low birth weight is considered a marker of social risk related to precarious socioeconomic conditions and maternal behavior in relation to health care.

Moreover, low birth weight can be understood as a sentinel event for healthcare services, which indicates the low quality of prenatal care and the need for training the staff in order to improve the identification of and the care provided to these groups of patients. Other actions such as increased access to prenatal care, compliance with protocols, and use of the proper criteria for high-risk pregnancies, suggested by the Ministry of Health, could directly reduce low birth weight and low Apgar scores (Gaíva et al., 2013).

Malformation can be caused by genetic and environments factors (e. g. use of alcohol and tobacco during the pregnancy) (Mendes et al., 2018). This information is relevant because some congenital genetic, infectious, or environmental-related anomalies can be prevented through the implementation of public policies and an adequate offer of health services.

In turn, prenatal care represents the most important protection for neonatal and infant survival, a situation confirmed by this study. These findings are somewhat in line with other research that focused on an insufficient number of prenatal visits and increased neonatal deaths (Nascimento et al., 2008). In the present study, 70.7% of the mothers that lost their babies made less than 7 prenatal appointments.

Prematurity (less than 37 weeks) are recognized as relevant factors for infant death, especially early neonatal death (Victora et al., 2001; Martins et al., 2004; Ortiz and Oushiro, 2008; Santos, 2012; Gaiva et al., 2014). These aspects are directly related with maternal conditions and prenatal care, which, in turn, work on several determinants and conditions of infant mortality, potentially reducible due adequate prenatal care (Ortiz and Oushiro, 2008; Santos, 2012; Gaiva et al., 2014).

The prenatal and biological attribute were key determinants. Despite the previous evidence in the literature, his study found no importance between neonatal mortality and maternal profile. This finding reinforces the importance of an adequate surveillance of deliveries and qualified care addressed to the newborn as a way to reduce infant morbimortality.

## Conclusions

Along with this paper, we proposed a new method to expose neonatal death risk-based in a combination of machine learning classifiers and demographic features. From public data collected from the Brazilian government, we created new datasets, comprising more than 30 million samples for the problem of neonatal mortality.

Furthermore, on features distribution on death samples had been performed, obtaining results which lead us to conclusions as the low influence of sex in neonatal mortality between 2006 and 2016, on median days of life (until death) in Brazil.

With results exceeding 85% AUC when using XGBoost as a final classifier, the method is able to provide both a death risk response and an interpretation of the result obtained. Between results using three different machine learning classifiers with their default parameters, points toward expressiveness of features, being newborn weight, Apgar at the fifth minute, congenital malformations, Apgar at the first minute, gestational weeks and number of prenatal appointments the six more expressive, respectively.

As a decision support tool, this kind of method can be useful to help health experts to take decisions if more intensive care is necessary for newborns in Brazil.

Additionally, from a demographic point of view, studies based on data analysis are valuable to corroborate important statements, once most of the studies are performed in small populations without an expressive statistical sample. Therefore, the findings of this study confirm our initial hypothesis.

The present study underscores the importance of cohort studies, which constitute an essential tool for the monitoring, evaluation and discussion on the health. For future research directions, our research intends to evaluate new methods for dealing with data encoding, such as categorical embeddings, as well as combinations between different classifiers in order to increase positive class (death) accuracy, for the occurrence of false negatives is a very problematic issue on methods related with health and perform same analysis over data extracted for all cities in Brazil.

## Data Availability

The dataset used in this paper is available.

## Ethics

This paper uses publicly available data (SIM and SINASC) that has been de-identified and was deemed exempt from human subjects review.

## References

1. Barbieri M. Infant and child mortality in the less developed world. International Encyclopedia of the Social & Behavioral Sciences 2015; 12: 21–26.

2. Billari FC, Fürnkranz J, Prskawetz A. Timing, sequencing, and quantum of life course events: a machine learning approach. European Journal of Population 2006; 22: 37–65.

3. Breiman L. Random forests. Machine learning 2001; 45(1): 5-32.

4. Chen T, Guestrin C. Xgboost: A scalable tree boosting system. In: Proceedings of the 22nd acm sigkdd international conference on knowledge discovery and data mining. ACM, 2016, pp. 785–794.

5. Chung SH, Choi YS, Bae CW. Changes in the neonatal and infant mortality rate and the causes of death in Korea. Korean Journal of Pediatrics 2011; 54(11):443–455.

6. Cortes C, Vapnik V. Support-vector networks. Machine learning 1995; 20(3): 273-297.

7. De Rose A, Pallara A. Survival trees: an alternative non-parametric multivariate technique for life history analysis. European Journal of Population 1997; 13: 223–241.

8. Duarte JLMB, Mendonça GAS. Factors associated with neonatal mortality among very low birthweight newborns in four maternity hospitals in the city of Rio de Janeiro, Brazil. Cad. Saúde Pública 2005; 21(1):181–191.

9. França E, Lansky S. Mortalidade infantil e neonatal no Brasil: situação, tendências e perspectivas. Rede Interagencial de Informações para Saúde-Demografia e Saúde: contribuição para análise de situação e tendências. Série Informe de Situação e Tendências 2009, p.83–112.

10. França E, Lansky S [homepage on the Internet]. Mortalidade infantil neonatal no Brasil: situação, tendências e perspectivas [cited 2016 Jun 15]. Available from: http://www.abep.nepo.unicamp.br/encontro2008/docsPDF/ABEP2008_1956.pdf

11. Gaíva MAM, Bittencourt RM, Fujimori E. Early and late neonatal death: characteristics of mothers and newborn. Rev Gaúcha Enferm 2013;34(4):91–97.

12. Gaíva MA, Fujimori E, Sato AP. Neonatal mortality in infants with low birth weight. Rev Esc Enferm USP 2014; 48: 778-86.

13. Garcia LP, Fernandes CM, Traebert J. Risk factors for neonatal death in the capital city with the lowest infant mortality rate in Brazil. J Pediatr 2019; 95: 194-200.

14. Hsieh MH, Hsieh MJ, Chen CM, Hsieh CC, Chao CM, Lai CC. Comparison of machine learning models for the prediction of mortality of patients with unplanned extubation in intensive care units. Scientific Reports 2018; 8: 2045-2322.

15. Instituto Brasileiro de Geografia e Estatística. Tábuas de mortalidade. 2010. http://www.ibge.gov.br/home/estatistica/populacao/projecao_da_populacao/2013/

16. Kuhn M, Johnson K. Applied predictive modeling. New York, NY: Springer, 2013.

17. Lima EFA, Sousa AI, Griep RH, Primo CC. Risk factors for neonatal mortality in the city of Serra, Espírito Santo. Rev Bras Enferm 2012; 65(4): 578-85.

18. Machado J, Cotta R, Soares J. Reflexões sobre o processo de municipalização das políticas de saúde: a questão da descontinuidade político-administrativa. Interface Comun Saude Educ 2015; 19:159–170, 2015.

19. Martins EF, Velásquez-Meléndez G. Determinants of neonatal mortality in a cohort of born alive infants, Montes Claros, Minas Gerais, 1997–1999. Rev Bras Saúde Matern Infant 2004; 4: 405-12.

20. Mendes IC, Jesuino RSA, Pinheiro DS, Rebelo ACS. Congenital anomalies and its main avoidable causes: a review. Rev Med Minas Gerais 2018; 28: e-1977. DOI: http://dx.doi.org/10.5935/2238-3182.20180011

21. Migoto MT, Oliveira RP, Silva AMR, Freire MHS. Early neonatal mortality and risk factors: a case-control study in Paraná state. Revista Brasileira de Enfermagem 2018; 71: 2527-2534.

22. Ministério da Saúde/Ministry of Health. Síntese de evidências para políticas de saúde: reduzindo a mortalidade perinatal. Departamento de Ciência e Tecnologia, Brasília: Ministério da Saúde, 2015. 43p.

23. Mosley W, Chen L. An analytical framework for the study of child survival in developing countries. Bulletin World Health Organization 2003; 81: 140-145.

24. Nascimento EMR, Costa MCN, Mota ELA, Paim JS. Investigation of risk factors for infant mortality by linking health databases. Cad. Saúde Pública 2008; 24: 2593-2602.

25. Nascimento RM, Leite AJM, Almeida NMGS, Almeida PC, Silva CF. Determinantes da mortalidade neonatal: estudo caso-controle em Fortaleza, Ceará, Brasil. Cadernos de Saúde Pública 2012; 28: 559-572.

26. Nguyen G. Evaluating statistical and machine learning methods to predict risk of in-hospital child mortality in Uganda., Dissertation (Master)—Public Health, 2016.

27. Ortiz LP, Oushiro DA. Profile of the neonatal mortality in the State of Sao Paulo. São Paulo Perspec 2008; 22: 19-29.

28. Palloni A, Pinto-Aguirre G. Adult mortality in Latin America and the Caribbean. In: Rogers R G, Crimmins EM(ed.). International handbook of adult mortality. New York, NY: Springer, 2011. p. 101–132.

29. Pan I, Nolan LB, Brown RR, Khan R, van der Boor P, Harris DG, Ghani R. Machine learning for social services: a study of prenatal case management in Illinois. American Journal of Public Health 2017; 107: 938-944.

30. Podda M, Bacciu D, Micheli A, Bellu R, Placidi G, Gagliardi L. A machine learning approach to estimating preterm infants survival: development of the preterm infants survival assessment (pisa) predictor. Scientific Reports 2018; 8: 2045-2322.

31. Prati RC, Batista GE, Monard MC. Data mining with imbalanced class distributions: concepts and methods. In: IICAI, 2009, pp. 359–376.

32. Santos HG. Fatores de risco para mortalidade Infantil em Londrina (PR): análise hierarquizada em duas coortes de nascidos vivos [master’s thesis]. Londrina (PR): UEL, 2012.

33. Singha AK et al. Application of machine learning in analysis of infant mortality and its factors. Working Paper, India, p. 1–5, 2016.

34. UNICEF. Committing to Child Survival: A Promise Renewed—Progress Report 2015. 96p. https://www.unicef.org/publications/index83078.html

35. Vasconcelos AMN, Gomes MMF. Transição demográfica: a experiência brasileira. Epidemiologia e Serviços de Saúde 2012; 21: 539-548.

36. Victora CG, Barros FC. Infant mortality due to perinatal causes in Brazil: trends, regional patterns and possible interventions. Sao Paulo Med J 2001; 119(1):33–42.

37. Waldemar CA, Shivaprasad SG, Imtiaz FCPS, Elwyn C, Antoinette T, Garces A, et al. Newborn-care training and perinatal mortality in developing countries. N Engl J Med 2010;362(7):614–23. Available from: http://www.nejm.org/doi/pdf/10.1056/NEJMsa0806033.

38. World Health Organization. Global reference list of 100 core health indicators (plus health-related SDGs 2018). World Health Organization, 2018. 162p.

